# Dual-stage AI system for Pathologist-Free Tumor Detection and subtyping in Oral Squamous Cell Carcinoma

**DOI:** 10.1101/2025.06.05.25329090

**Authors:** Nisha Chaudhary, Prateeksha Muddemanavar, Deepak Kumar Singh, Arpita Rai, Deepika Mishra, SV Sowmya, Dominic Augustine, Jeyaseelan Augustine, Akhilesh Chandra, Akhilanand Chaurasia, Tanveer Ahmad

## Abstract

**Background:** Accurate histological grading of oral squamous cell carcinoma (OSCC) is critical for prognosis and treatment planning. Current methods lack automation for OSCC detection, subtyping, and differentiation from high-risk pre-malignant conditions like oral submucous fibrosis (OSMF). Further, analysis of whole-slide image (WSI) analysis is time-consuming and variable, limiting consistency. We present a clinically relevant deep learning framework that leverages weakly supervised learning and attention-based multiple instance learning (MIL) to enable automated OSCC grading and early prediction of malignant transformation from OSMF.

**Methods:** We conducted a multi-institutional retrospective cohort study using a curated dataset of 1,925 whole-slide images (WSIs), including 1,586 OSCC cases stratified into well-, moderately-, and poorly-differentiated subtypes (WD, MD, and PD), 128 normal controls, and 211 OSMF and OSMF with OSCC cases. We developed a two-stage deep learning pipeline named *OralPatho*. In stage one, an attention-based multiple instance learning (MIL) model was trained to perform binary classification (normal vs OSCC). In stage two, a gated attention mechanism with top-K patch selection was employed to classify the OSCC subtypes. Model performance was assessed using stratified 3-fold cross-validation and external validation on an independent dataset.

**Findings:** The binary classifier demonstrated robust performance with a mean F1-score exceeding 0.93 across all validation folds. The multiclass model achieved consistent macro-F1 scores of 0.72, 0.70, and 0.68, along with AUCs of 0.79 for WD, 0.71 for MD, and 0.61 for PD OSCC subtypes. Model generalizability was validated using an independent external dataset. Attention maps reliably highlighted clinically relevant histological features, supporting the system’s interpretability and diagnostic alignment with expert pathological assessment.

**Interpretation:** This study demonstrates the feasibility of attention-based, weakly supervised learning for accurate OSCC grading from whole-slide images. *OralPatho* combines high diagnostic performance with real-time interpretability, making it a scalable solution for both advanced pathology labs and resource-limited settings.

## Introduction

OSCC is one of the most prevalent malignancies worldwide and represents a major public health burden, especially in South and Southeast Asia, where it ranks among the leading causes of cancer-related morbidity and mortality (1). Despite advancements in surgery, radiotherapy, and immunotherapy, clinical outcomes for OSCC remain suboptimal due to late-stage diagnosis, limited access to expert histopathological services, and significant inter-observer variability in tumor grading (2,3). Histological classification into WD; MD; and PD subtypes forms a cornerstone for determining prognosis and guiding therapy (4,5). However, this task is inherently subjective and labour-intensive, making it an ideal candidate for augmentation using artificial intelligence (AI).

In recent years, deep learning methods have shown remarkable potential in automating cancer diagnosis and subtyping from histopathology images (6–11). Particularly in breast and lung cancer, AI systems trained on WSIs have achieved high diagnostic accuracy and have demonstrated clinical relevance. For instance, (12) used convolutional neural networks (CNNs) to classify lung adenocarcinoma subtypes and predict key driver mutations directly from WSIs, while (13) applied weakly supervised learning across multiple cancers, including prostate and breast, in a clinical-grade framework. Similarly, (14) and (15) developed deep learning-based pipelines for breast cancer subtyping and metastasis detection in lymph nodes, respectively, offering insights into both performance and interpretability. These landmark studies have laid the groundwork for computational histopathology, but there remains a significant gap in applying similar rigor and scalability to oral cancer.

Previous studies on OSCC have predominantly focused on binary classification, which helps in distinguishing tumor from normal tissue using CNNs and transfer learning approaches (16–19). For example, (17) reported high classification accuracy using transfer learning-based model; however, their framework lacked multiclass grading capability and offered limited interpretability. Similarly, (20,21) applied CNN-based classifiers for OSCC detection but did not incorporate attention mechanisms or assess generalizability across diverse clinical datasets. Notably, many of these efforts were constrained by small, single-institution datasets, which restrict scalability and hinder real-world clinical translation. The lack of large, well-annotated datasets for OSCC sub typing remains a significant barrier to developing robust AI-based diagnostic tools.

Furthermore, most existing models operate as black boxes, offering no visual explanation for their predictions, a critical limitation when trust, interpretability, and transparency are prerequisites for clinical adoption. To help overcome data scarcity and enhance training quality, we recently introduced the ORCHID (Oral Cancer Histopathology Imaging Dataset), which is an open-access repository comprising digitized oral cancer images captured at 100x magnification (22). ORCHID enables detailed visualization of nuclear and tissue-level features essential for OSCC grading and represents one of the first standardized high-resolution datasets in this domain. However, it currently lacks low-resolution whole-slide context, which limits its suitability for multi-scale analysis. Moreover, annotations are confined to image-level labels without region-specific segmentation or multi-expert consensus, thereby limiting its immediate application in supervised patch-level training and explainability benchmarking.

To address existing limitations in OSCC diagnosis, we present *OralPatho*, a dual-stage, attention-based AI framework for automated and interpretable grading of OSCC from WSIs. Leveraging weakly supervised multiple instance learning (MIL) with gated top-K patch selection, *OralPatho* performs both binary classification (normal vs tumor) and multiclass subtyping (WD, MD, PD) without the need for detailed annotations. The model is trained on the largest known multi-institutional OSCC dataset to date 1,586 WSIs from five diverse Indian centers, ensuring robust generalizability across staining protocols and populations.

*OralPatho* offers clinically meaningful interpretability through attention heatmaps that localize key histological features and a browser-based interface for real-time slide upload, visualization, and inference. By combining scalability, explainability, and deployment readiness, *OralPatho* extends MIL-based digital pathology to an underserved cancer type and lays the groundwork for future integration with multimodal data and adaptive learning for precision oncology.

## Methods

### Data sources and cohorts

This retrospective, multi-institutional study was designed to develop and validate an AI-based diagnostic framework for OSCC using digitized histopathology slides. A total of 813 patient histopathology data were retrospectively collected from six leading medical institutions in India, All India Institute of Medical Sciences (AIIMS), Banaras Hindu University (BHU), Jamia Millia Islamia (JMI), Maulana Azad Institute of Dental Sciences (MAIDS), Rajendra Institute of Medical Sciences (RIMS), and Ramaiah University of Applied Sciences (RUAS) along with publicly available normal and SCC slides from The Cancer Imaging Archive (TCIA-https://www.cancerimagingarchive.net/) for oral cancer. The dataset for this study comprised 459 tissue slides of OSCC (260 from TCIA), 111 OSMF, 100 OSMF-OSCC and 128 normal tissue condition (105 from TCIA, 9 from RIMS and 13 JMI). Patient demographic information including age, gender, and lesion site was aggregated across cohorts (see Supplementary **Figure SF3**)

Each physical glass slide was digitized into one or more WSIs depending on the number of spatially distinct tissue regions, yielding a total of 1,586 WSIs for downstream analysis. Expert pathologists annotated each slide with one of three OSCC grades; WD, MD, or PD, based on consensus histological criteria. No patch-level annotations were used during training.

All slides were anonymized and de-identified, and data collection was conducted under institutional review board (IRB) approvals at each contributing site.

Throughout this study, the section-level WSI count shown in **Figure 1A** is used as the official dataset reference, reflecting the effective number of spatially distinct tissue regions analyzed. This distinction accounts for the fact that a single glass slide may contain multiple spatially separated tissue sections, each processed as an independent WSI for AI model development and evaluation (**Figure SF7**).

**Figure 1:**
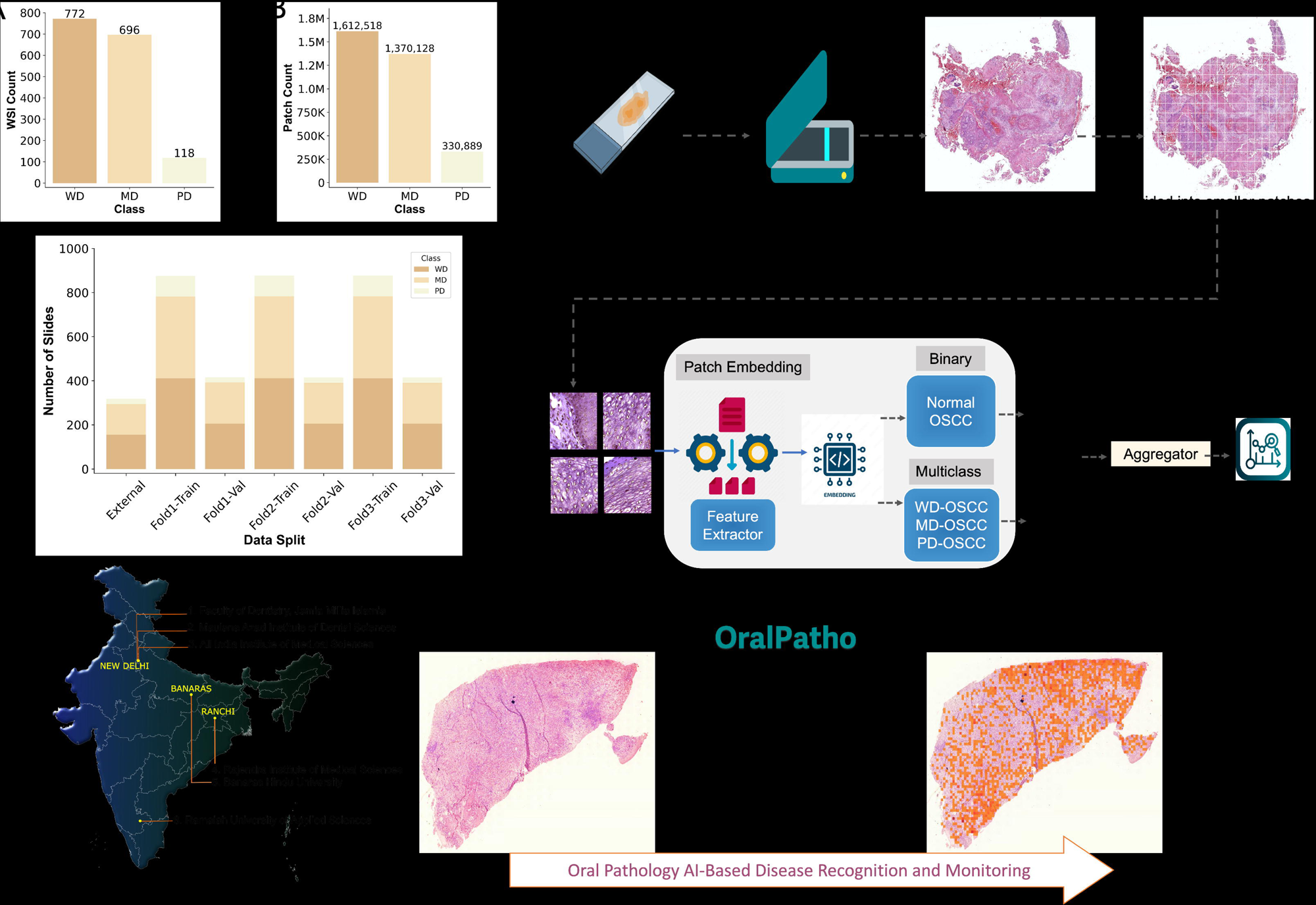
Cohort Summary, Data Distribution, and System Overview of *OralPatho* Platform. A. Total number of annotated Whole Slide Images (WSIs) per class — well-differentiated (WD), moderately differentiated (MD), and poorly differentiated (PD) oral squamous cell carcinoma (OSCC). B. Tumor patch counts extracted from WSIs across classes, reflecting the scale of patch-level supervision. C. Distribution of WSIs across external test and 3-fold cross-validation sets, stratified by histologic subtype. D. Geographic provenance of patient data across five major medical and dental institutions in India, capturing a diverse and representative OSCC cohort. E. Overview of the diagnostic pipeline from tissue slide acquisition, digitization, patch-level segmentation, and feature extraction to dual-stream MIL classification. Binary classification distinguishes OSCC from normal, while multiclass classification further stratifies OSCC into WD, MD, and PD subtypes. F. Interface snapshot from the *OralPatho* platform, showcasing representative histology slides and output prediction visualization for AI-assisted monitoring and diagnosis.

### WSI processing and patch extraction

WSIs were divided into non-overlapping patches of 256 × 256 pixels, extracted from the highest resolution level of the WSI, with their coordinates saved, ensuring the capture of localized histopathological patterns and structures. Different libraries, including OpenSlide for WSI handling, skimage, scipy, and cv2 for image processing like Canny edge detection, were used. A tissue threshold of 0.75 was applied to select patches with at least 75% tissue area, focusing on diagnostically relevant areas and excluding those with insufficient tissue content. Colour normalization(detailed in supplementary methods) was applied to minimize staining variability across sources, and data augmentation including random rotations, flips, and contrast adjustments was performed to enhance model generalizability. Extracted patches were processed into a structured dataset of HDF5 files, containing both the image patches and associated metadata for downstream processing. Each WSI was subjected to preprocessing, including background segmentation and patch generation.

### AI model architecture and training overview

Patch-level feature embeddings were extracted using a convolutional neural network (CNN), adapted for histopathology image analysis. ResNet50, modified for feature extraction, was our main choice because of its strong residual connections, which effectively avoid vanishing gradient problems while capturing complex histopathological features. Although DenseNet201 and EfficientNet were tested, they were not prioritized because of their additional computational complexity and no significant improvements for our specific OSCC dataset.

We implemented a two-stage attention-based multiple instance learning (MIL) framework. In the first stage, a binary MIL model identified OSCC vs. normal WSIs. In the second stage, a multiclass MIL model classified OSCC into well-, moderately-, or poorly-differentiated subtypes. Both stages used 512-dimensional patch-level features extracted by a ResNet50 backbone. An attention mechanism was applied to assign importance weights to patches, enabling the model to focus on the most diagnostically relevant regions. A top-K pooling strategy was used to retain high-attention patches for final slide-level predictions (**Figure 2**). Model training was conducted using stratified three-fold cross-validation and an independent held-out test set. The Adam optimizer with a learning rate of 1e-4 and batch size of 64 was used, with early stopping based on validation loss to prevent overfitting (see Supplementary **Table S1**).

**Figure 2:**
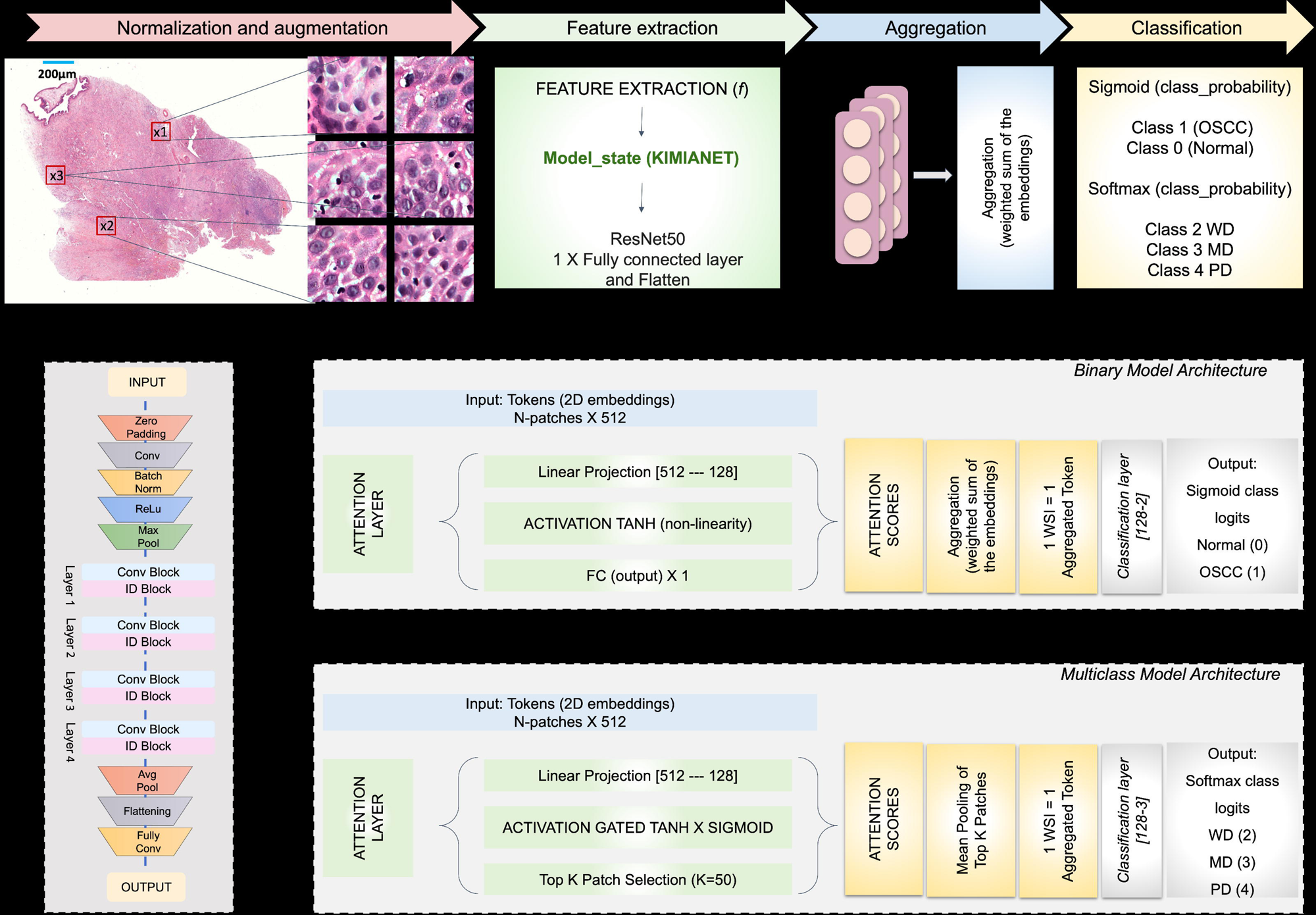
Model Pipeline and Architecture for Patch-level OSCC Classification. A. End-to-end pipeline overview illustrating slide normalization, patch-level sampling and augmentation, feature extraction using a ResNet50-based encoder (termed KIMIANET), and subsequent classification using MIL-based aggregation. Binary and multiclass branches diverge at the final classifier stage. B. Schematic of the modified ResNet50 architecture used to generate 512-dimensional embeddings from histology patches. The network contains convolutional blocks interleaved with identity (ID) blocks for stable feature extraction. C. Binary attention-based MIL model architecture: each patch embedding is processed through an attention layer with Tanh non-linearity to generate attention scores, which are then used to compute a weighted average across patches. The aggregated slide-level representation is passed through a classification layer yielding a binary label-Normal (0) or OSCC (1). D. Multiclass MIL model with Top-K patch selection: similar attention formulation but includes a Gated-Tanh-Sigmoid activation and retains only the top 50 most informative patches. Their mean-pooled representation is classified into one of three OSCC grades WD, MD, or PD.

### Interface Development and Deployment

We developed a lightweight web-based interface to enable clinical translation of our model. The platform supports WSI upload, real-time OSCC grading, and patch-level attention visualisation. The interface was built using a modular architecture compatible with standard digital pathology formats (e.g., .svs, .tiff). Designed for low-latency operation, the system enables deployment in remote and resource-limited settings where access to expert pathology may be constrained.

## Results

### Multi-institutional dataset design and interactive AI workflow for OSCC grading

A large, multi-institutional dataset comprising of 1,586 OSCC WSIs, was curated and categorized into WD, MD, and PD subtypes, as shown in **Figure 1A**, followed by expert pathologists annotation of each slide. The dataset includes 772 WD slides (∼1.6M tumor patches), 696 MD slides (∼1.37M patches), and 118 PD slides (∼0.33M patches) (**Figure 1B**), revealing significant class imbalance, with PD cases comprising only 7.4%. To mitigate this and ensure robust evaluation, WSIs were stratified by subtype and distributed across three cross-validation folds and an external test set (**Figure 1C**). Samples were sourced from institutions across India, introducing variation in staining, scanner types, and patient demographics (**Figure 1D**), enhancing model generalizability and better reflecting real-world pathology workflows.

**Figure 1E** shows representative histological features including keratin pearls, nuclear pleomorphism, and epithelial disarray, which are used by pathologists and targeted by our model’s attention heatmaps. To operationalize this, we developed the *OralPatho* workflow and an interactive browser-based interface (**Figure 1F**), enabling WSI upload, real-time prediction, and patch-wise heatmap visualization. Together, these components demonstrate the clinical relevance, interpretability, and deployment readiness of the *OralPatho* platform.

### Architecture and workflow of the dual-stage *OralPatho* framework for weakly supervised OSCC detection and grading

The *OralPatho* model architecture illustrates the complete computational workflow employed for both binary tumor detection and OSCC subtype classification, as detailed in **Figure 2**. The pipeline begins with slide normalization and patch-level extraction, where WSIs are divided into 256×256 non-overlapping patches following normalization (**Figure 2A**). Each patch is encoded into a fixed-length 512-dimensional vector using a modified ResNet50 architecture (**Figure 2B**). This architecture, built on the standard ResNet50 framework, integrates convolutional and identity blocks to capture both local and contextual histological features. The modifications include replacing the average pooling layer with an adaptive average pooling layer and replacing the fully connected layer with a linear layer mapping 2048 dimensions to 512, followed by a ReLU activation. The binary classification task (normal vs tumor) is handled by MIL framework (**Figure 2C**), in which each patch embedding is passed through a Tanh-based attention module to compute importance weights. These weights are aggregated to produce a slide-level representation, which is then classified into normal or OSCC. For subtype grading, the model extends this architecture with a gated attention mechanism that combines Tanh and Softmax activations to refine attention scores (**Figure 2D**). A top-K patch selection strategy retains the 50 most informative patches per slide based on validation performance, whose embeddings are mean-pooled and used to classify the slide into one of three histological grades: WD, MD, or PD. This dual-stage architecture reflects a careful balance between scalability, diagnostic accuracy, and interpretability. By leveraging weak supervision, the model learns effectively from slide-level labels without requiring region-specific annotations. The attention-based design and top-K selection enhance both performance and transparency, focusing the model on diagnostically relevant regions in a manner aligned with expert pathologist review.

### Quantitative accuracy and visual explainability of the *OralPatho* framework for OSCC grading

The confusion matrices for the binary classification task (normal vs. OSCC) across three cross-validation folds are shown in **Figure 3A**. The model demonstrated consistently high performance, with F1-scores exceeding 0.95, indicating robust differentiation between tumor and normal tissue across heterogeneous WSIs. Detailed per-class performance across three folds is provided in Supplementary **Table S3**. The low rate of misclassification and strong row-wise consistency in predictions confirm the reliability of our attention-based MIL framework in primary tumor screening. We analyzed the performance of our multiclass MIL model in classifying OSCC subtypes into the three categories and achieved a macro-averaged F1-score of 0.84 ± 0.02, with WD and MD subtypes classified accurately across all folds (**Figure 3B**). Although PD samples showed a moderate decline in precision and recall, we attribute this to their lower representation and increased morphological heterogeneity. Nonetheless, the model consistently maintained subtype-specific discrimination, reinforcing its clinical reliability(Supplementary **Table S4**).

**Figure 3:**
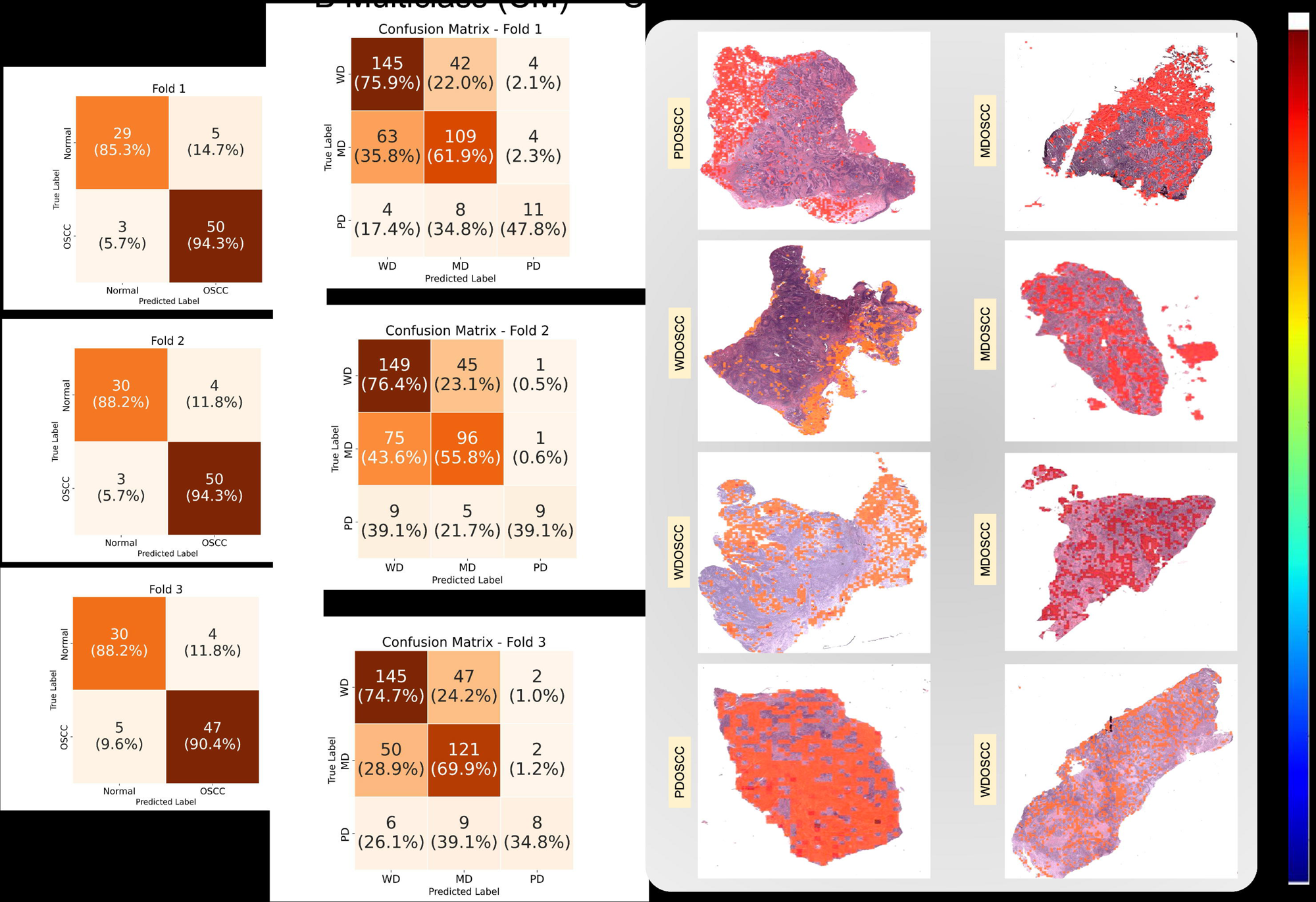
Evaluation of Model Performance and Interpretability via Heatmaps. A. Confusion matrices for the binary classification model across 3 folds. Values inside cells indicate both count and row-wise prediction accuracy (as percentages). B. Confusion matrices for the multiclass classification model (WD, MD, PD) across the same folds. Each matrix reflects the model’s ability to distinguish histological subtypes of OSCC, with clear class-wise performance disparities. C. Heatmaps overlaid on whole-slide images showing attention-based patch-level predictions from the model. Redder regions correspond to higher attention weights, highlighting regions contributing most strongly to the model’s decision-making. Each example is labeled with its true OSCC subtype, demonstrating consistent attention localization across all three histopathological classes.

To investigate interpretability, attention-based heatmaps were visualized (**Figure 3C**) by overlaying model-derived attention scores onto the original WSIs. Red-highlighted regions indicate areas of high diagnostic relevance as identified by the model. The attention maps reliably localized key pathological features without requiring pixel-level supervision, highlighting keratin pearls in WD, cellular disorganization in MD, and nuclear atypia in PD OSCC. Occasional mislocalizations were observed in regions with dense inflammation or artefacts, primarily affecting PD predictions. Overall, the model exhibited strong spatial focus and interpretability aligned with expert histopathological criteria.

To validate the generalizability of the attention mechanisms, evaluation was extended to external datasets, as shown in **Figure SF1-A**. The model was applied to institutional OSCC slides not encountered during training, and the resulting heatmaps confirmed accurate localization of tumor-rich regions, indicating strong intra-institutional consistency. Further assessment using publicly available OSCC WSIs from the TCIA database is shown in **Figure SF1-B**. Despite differences in staining protocols and imaging systems, the model maintained robust attention localization, consistently focusing on diagnostically relevant tissue regions. The color gradients from blue (low attention) to red (high attention) visually confirmed the transferability of our learned feature representations. Together, these results demonstrate that our dual-stage attention-based framework not only performs robustly across diverse cohorts but also generates biologically interpretable outputs.

### External validation of *OralPatho*: classification performance and latent feature visualization

The generalizability and robustness of the *OralPatho* multiclass classification framework were assessed using an independent external test cohort sourced from multiple institutions and scanner types (**Supplementary Table S2**). Confusion matrices generated for each cross-validation fold (**Figure 4A**) demonstrated consistent class-wise prediction performance for WD, MD, and PD OSCC subtypes. The model maintained high predictive accuracy across all folds, with row-normalized metrics reflecting stability on real-world, unseen data. WD and MD subtypes were classified with high accuracy, while PD cases, though more challenging due to class imbalance and morphological heterogeneity, were still detected with reasonable sensitivity. **Figure SF2** further illustrates accurate PD detection despite its low representation and structural ambiguity.

**Figure 4:**
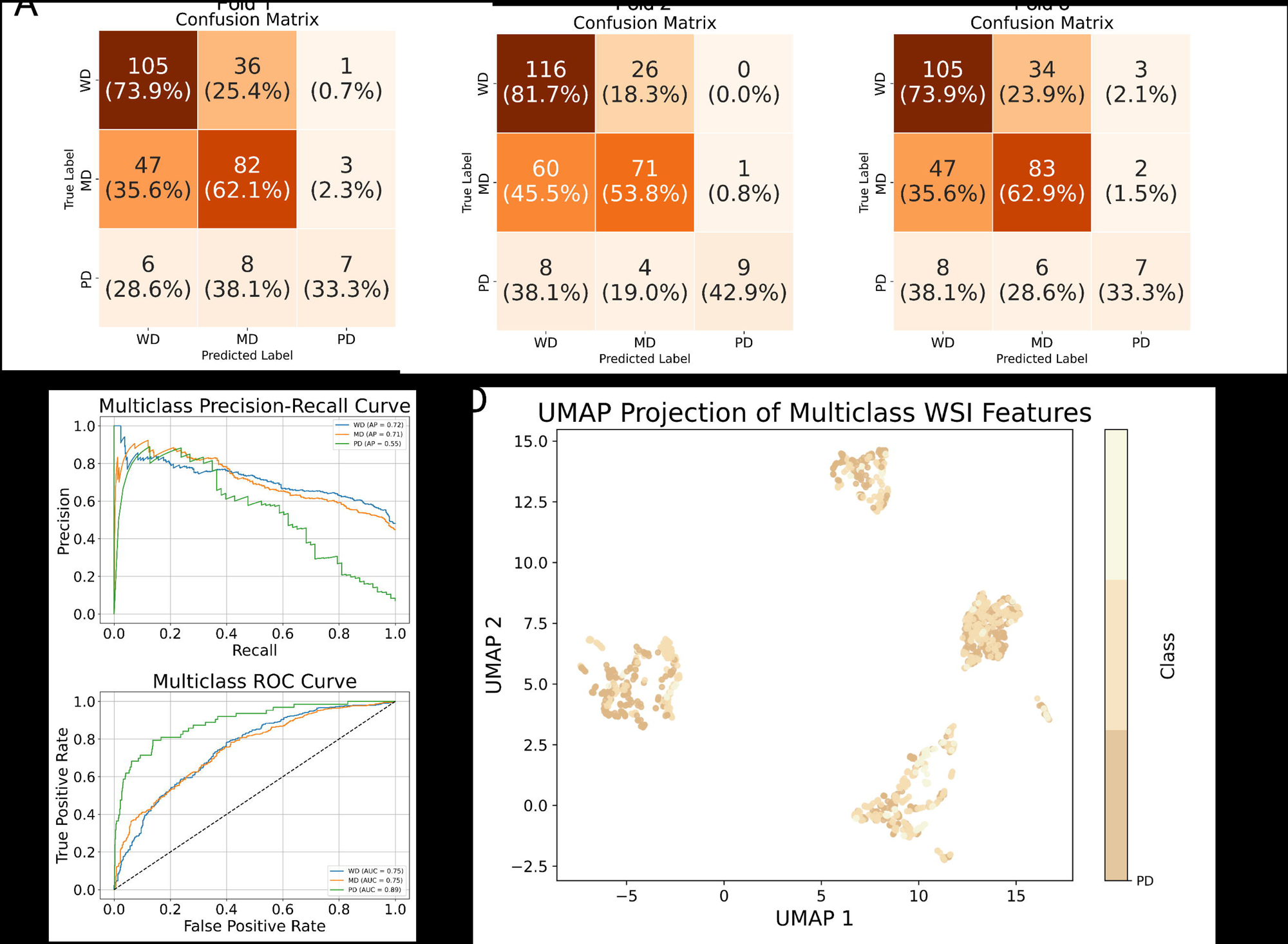
Performance of the Multiclass Model on External Test Cohort. A. Confusion matrices for each fold on external test slides (n = 318), with class-wise counts and row-normalized prediction accuracy. B. Precision-recall curves for each OSCC subtype (WD, MD, PD) computed using the fused predictions across all folds. C. Multiclass ROC curves reflecting overall classification discrimination. Area under the curve (AUC) is reported for each subtype, showing distinct prediction capability per class. D. UMAP visualization of the learned WSI-level features from the external dataset. Slides are color-coded by histological subtype, demonstrating how the model encodes subtype-specific representation in feature space.

To quantify subtype-level performance, we computed precision-recall curves for each class (**Figure 4B**). WD and MD achieved high precision and recall, while the PD curve, although moderate, retained sufficient sensitivity to reflect meaningful differentiation. Notably, the model avoided collapsing into majority class predictions, consistently preserving discriminative capability across all histological types. Receiver operating characteristic (ROC) curves (**Figure 4C**) further supported these findings, yielding AUC scores of 0.871 (WD), 0.823 (MD), and 0.731 (PD), confirming strong subtype separability and reliable classification even in the presence of cross-institutional variability in tissue preparation, staining, and imaging. To explore the model’s learned feature space, we generated a UMAP of WSI-level embeddings from the external test set (**Figure 4D**). Slides were color-coded by subtype, revealing distinct clustering patterns that aligned with clinical expectations. WD and PD cases formed well-separated clusters, while MD cases were more diffusely distributed along a transitional continuum, reflecting their histological intermediate nature. This latent space organization demonstrates that the model’s internal representations capture biologically meaningful patterns and support subtype-specific decision-making. Together, these results validate *OralPatho*’s cross-site robustness, diagnostic precision, and explainable internal representations, emphasizing its suitability for deployment in diverse clinical and resource-limited settings.

### Model reveals variable detection confidence in transitional OSMF and OSMF-OSCC lesions

To evaluate the model’s behavior on pre-malignant lesions, we applied the trained *OralPatho* framework to a held-out set of 111 only OSMF WSIs, which were not seen during training. As shown in **Figure 5A**, although the model was not trained on OSMF, several slides received OSCC probabilities in the range of 0.27-0.30. These cases represent histological overlap between benign and early malignant patterns. **Figure 5B** presents representative visualizations for the top-ranking OSMF slides, including the raw WSI, model attention overlay, and high-magnification crops from regions of interest. The attention maps frequently localized to the basal epithelium and regions showing nuclear crowding or architectural irregularity, demonstrating the model’s ability to selectively focus on plausible high-risk areas without overcalling noise. This highlights the potential utility of *OralPatho* not only for diagnostic classification but also for risk stratification in ambiguous pre-cancerous states. We further extended the analysis to a distinct held-out set of 100 OSMF-OSCC cases, representing transitional lesions with confirmed malignant transformation under a background of OSMF. As shown in Supplementary **Figure SF4**, model-generated attention heatmaps varied substantially across these slides. While some (e.g., Cases 1-5) exhibited moderate to high attention in areas showing epithelial dysplasia or early invasion, others (Cases 6-10) were largely ignored by the model despite containing subtle cues. Expert pathologist review (**Table S5**) estimated region-level attention accuracy to range from 20% to 75%, with higher correspondence in slides containing early invasive foci. Quantitatively, as shown in Supplementary **Figure SF5,6**, the mean attention scores for OSMF and OSMF-OSCC slides were significantly lower than those for well-differentiated OSCC (WDOSCC; p < 0.0001), moderately differentiated OSCC (MDOSCC; p < 0.01), and Normal slides (p < 0.0001). These findings reflect the model’s reduced confidence in histologically ambiguous samples and suggest its capacity to identify malignant cues in transitional lesions while avoiding overactivation in reactive or non-neoplastic areas. Together, this underscores *OralPatho*’s interpretability and conservative activation profile in borderline contexts.

**Figure 5:**
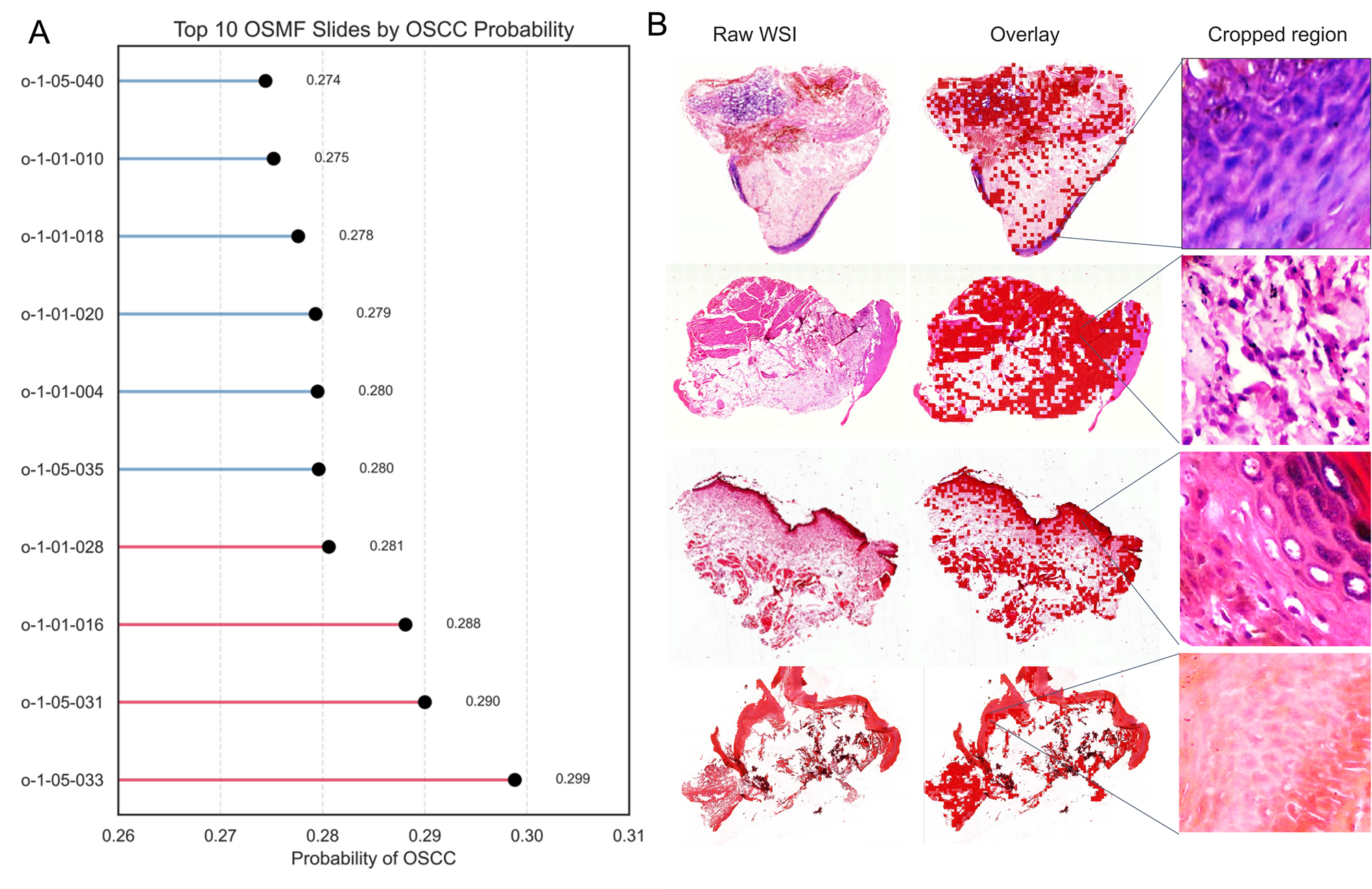
Interpretability analysis on OSMF slides. A. Confusion matrices for each fold on external test slides (n = 318), with class-wise counts and row-normalized prediction accuracy. B. Predicted OSCC probabilities for the top 10 OSMF slides using the trained binary model. For each top-ranked slide, the original WSI (left) and corresponding attention heatmap (right) are shown. High-attention regions mark areas the model deems OSCC-like. Notably, even without explicit training on OSMF, the model assigns localized importance to distinct epithelial zones in certain cases.

### Deployment-ready interface empowers real-time clinical decision-making

To support translational adoption, a modular, browser-accessible interface was developed, allowing clinicians to upload WSIs, perform automated OSCC grading, and visualize model-derived outputs in real time. The interface integrates slide-level predictions with attention-based patch overlays, allowing users to inspect high-attention regions corresponding to diagnostically relevant features **(Figure SF8)**. Designed for low-latency use, the interface supports batch uploads and is compatible with standard digital pathology formats. Visual feedback on tumour localisation and grading outcomes enhances user trust and provides a foundation for collaborative clinical validation or expert override when necessary. By combining robust classification with intuitive visual interpretability, this interface bridges the gap between research and practice especially in remote or under-resourced settings where access to trained pathologists may be limited. It represents a step toward scalable, AI-assisted decision support systems for histopathological evaluation of OSCC.

## Discussion

In this study, we present *OralPatho*, a clinically inspired, weakly supervised deep learning framework that performs automated, interpretable grading of OSCC from WSIs. The pipeline employs a two-stage architecture: first, an attention-based MIL model distinguishes tumor from normal tissue; second, a gated attention classifier with top-K patch selection enables fine-grained subtyping into WD, MD, and PD OSCC. This design eliminates the need for detailed region-level annotations and mimics a pathologist’s hierarchical decision-making approach by first confirming malignancy and then assigning a grade.

Consistent performance was observed across stratified three-fold cross-validation and an independent multi-institutional external test set, confirming the model’s robustness and generalizability across diverse data sources, staining protocols, and scanner types. These findings align with recent literature emphasizing the necessity of cross-site validation for AI models in digital pathology (13,14). *OralPatho* outperformed previous CNN-based OSCC classifiers (17,20) by integrating attention mechanisms and delivering subtype-specific predictions, which earlier models either did not attempt or failed to validate on external cohorts.

A critical strength of *OralPatho* is its interpretability. Attention heatmaps generated by the model consistently localized diagnostically relevant regions, including keratin pearls in WD, epithelial disarray in MD, and nuclear atypia in PD, without the need for pixel-level supervision. Such interpretability is essential for clinical trust and aligns with the growing emphasis on explainable AI in medicine (23,24). Additionally, our use of UMAP projections to visualize learned representations revealed a biologically meaningful separation of subtypes, further confirming the validity of the model’s internal logic.

While the model achieved high accuracy for WD and MD classifications, performance for PD cases was comparatively lower. This was primarily due to their underrepresentation (7.4%) and increased morphological variability, consistent with findings from related studies in breast and lung cancer histology where rare subtypes often challenge AI classification models (12,14). Furthermore, histopathological grading remains subjective, and the lack of multi-expert consensus annotations may have introduced label noise, especially in borderline or ambiguous cases. We also benchmarked model limitations explicitly, a practice often omitted in AI studies but critical for real-world deployment. Transparent analysis of failure modes in PD cases, particularly those misclassified due to dense inflammation or staining artifacts, reinforces the need for continual refinement. Integration of multimodal data, such as genomic markers or clinical metadata, could help disambiguate complex presentations and improve precision (25).

Importantly, the model had not been exposed to OSMF samples during training. As expected, the majority of OSMF cases were classified as “normal,” reflecting their non-malignant histological features. However, a small subset (10 WSIs) of OSMF cases were predicted to be as “OSCC”, suggesting that the model was able to identify subtle features reminiscent of malignancy in these slides. We further tested the model on OSMF-OSCC cases representing borderline lesions. Although excluded from training, the model localized high-attention regions in slides with clear malignant features, while others showed diffuse or low attention, mirroring diagnostic uncertainty. These results suggest potential utility in flagging ambiguous cases for closer review, though prospective validation is needed.

In addition to its core modelling contributions, *OralPatho* includes a lightweight, browser-based interface that supports slide upload, automated grading, and attention-based visualization. This deployment-ready platform bridges the gap between research and clinical use, offering a scalable solution suitable for both high-throughput laboratories and resource-limited healthcare environments where access to trained pathologists may be scarce. Looking ahead, we plan to expand the cohort diversity, enrich annotations through expert consensus, and explore continual learning and domain adaptation strategies to maintain performance across evolving clinical datasets. By combining technical rigor with interpretability and deployment readiness, *OralPatho* establishes a strong foundation for future AI-assisted diagnostic frameworks in head and neck oncology.

## Data Availability

The whole-slide images (WSIs) used in this study were obtained from institutional archives and public repositories. Due to patient privacy and institutional regulations, the institutional WSIs cannot be publicly shared. However, access may be granted for academic research purposes upon reasonable request and signing of a data use agreement. Publicly available WSIs from The Cancer Imaging Archive (TCIA) used in this study can be accessed at https://www.cancerimagingarchive.net/.

## Authors’ contributions

NC contributed to study conception and design, project administration, methodology, software development, data curation, formal analysis and interpretation, visualization, and drafting and critical revision of the manuscript. PM contributed to methodology, software development support, data curation, and revision of the manuscript. DKS contributed to figure preparation, reference curation, and manuscript reviewing. AR, DM, SSV, JA, AC, and DA contributed to data acquisition, pathological annotation, investigation, and manuscript review. AC and TA contributed to study supervision, project administration, resource provision, data validation, and manuscript editing and final approval. TA also provided overall study resources and coordinated multi-institutional collaboration. NC and TA accessed and verified the raw data and had full access to all study data. All authors reviewed the manuscript, contributed to critical revisions, and approved the final version for publication.

## Acknowledgments

We gratefully acknowledge the institutional support and computational infrastructure that enabled the successful execution of this study. This work was supported by the Science and Engineering Research Board (CRG/2020/002294) and the Indian Council of Medical Research (ICMR) (GIA/2019/000274/PRCGIA (Ver-1)), New Delhi, India. N.C. is the recipient of a Senior Research Fellowship from ICMR (3/1/2(1)/Oral/2021-NCD-II), which supported her research. We are grateful to our laboratory members, Faizan, Manisha, Divya, Varnit, Rohit, Kashif, Areej, Sufyan, Sheetal, Aashi, Ruquaiya, Shakir, Ubaid, and Nadeem, for their continuous support, constructive discussions, and encouragement throughout the course of this project. We also acknowledge the faculty and administrative staff at the Multidisciplinary Centre for Advanced Research and Studies (MCARS), Jamia Millia Islamia, for their continued encouragement and logistical assistance.

## Supplementary material

### Ethical approval

Ethical approvals for the collection and use of human tissue slides were obtained from the Institutional Ethics Committees (IECs) of all contributing institutions: Jamia Millia Islamia, New Delhi (Proposal No.: 6(25/7/241/JMI/IEC/2021); Maulana Azad Institute of Dental Sciences, New Delhi (Proposal No.: F./18/81/MAIDS/Ethical Committee/2016/8099); Rajendra Institute of Medical Sciences, Jharkhand (Proposal No.: ECR/769/INST/JH/2015/RR-18/236); Banaras Hindu University, Banaras (Proposal No.: Dean/2021/EC/2662); All India Institute of Medical Sciences, New Delhi (Proposal No.: IEC-828/03.12.2021, RP-33/2022) and Ramaiah University of Applied Sciences, Bangalore, India.

All samples were fully anonymized and de-identified prior to analysis to ensure privacy and confidentiality. This study includes the generation of new whole slide images (WSIs) from previously consented and ethically approved tissue samples, which had not been published in earlier datasets. No new tissue collection was conducted for the purposes of this work. The digital imaging, annotation, and AI model development processes adhered to the scope and stipulations of the original IRB approvals at each institution. Publicly available histopathology slides of OSCC and normal tissue were also included from The Cancer Imaging Archive (TCIA), which provides open-access anonymized data compliant with ethical use and re-distribution policies.

### Slide preparation and H&E staining

Formalin-fixed, paraffin-embedded (FFPE) tissue biopsies were sectioned at 4 μm thickness and mounted on standard glass slides. Hematoxylin and eosin (H&E) staining was performed according to institutional protocols, involving sequential immersion in hematoxylin, acid-alcohol differentiation, bluing reagent, and eosin. Slides were manually reviewed by certified pathologists to ensure diagnostic quality, appropriate staining contrast, and absence of artefacts (e.g., tissue folds, detachment). Only slides meeting these criteria were included for digitisation and downstream analysis. This process adhered to established guidelines for diagnostic H&E histopathology preparation (1).

(1) *Bancroft JD, Gamble M. Theory and Practice of Histological Techniques. 7th ed. Elsevier Health Sciences; 2008)*.

### Image scanning systems - Aperio, Hamamastu

Digitization was performed using Aperio AT2 (Leica Biosystems) and Hamamatsu NanoZoomer scanners across participating institutions. All slides were scanned at 40× magnification (0.25 µm/pixel). Scanner metadata, resolution levels, and file formats (.svs, .ndpi) were retained during preprocessing to ensure compatibility with downstream image extraction pipelines. Image handling and format standardization were managed using the OpenSlide library.

### Dataset preprocessing and de-identification protocol

The collected WSIs originated from heterogeneous sources with non-standardized file naming conventions. To enable scalable data handling, each WSI was programmatically renamed using a consistent format: o-<class_id>-<source_id>- <patient_id> and file extension (e.g., .svs or .ndpi). Class IDs mapped diagnostic categories (0 for normal, 1 for OSMF, 2 for WDOSCC, etc.), while source IDs identified contributing institutions (01: JMI, 02: MAIDS, etc.). The renaming was implemented using a Python script utilizing the os, pandas, and shutil libraries. This standardisation embedded diagnostic and source metadata into filenames, improving traceability and simplifying downstream processing workflows.

### Full WSI preprocessing & patch extraction protocol (size and filtering)

Whole-slide images (WSIs) were processed at the highest available resolution level (level-0; 0.25 μm per pixel) to ensure maximum spatial detail. Each WSI was divided into non-overlapping image tiles of 256×256 pixels using a fixed stride of 256 pixels across both spatial dimensions. A tissue detection strategy based on channel-wise Canny edge detection, followed by binary morphological operations (closing, dilation, and hole filling), was applied to exclude non-informative regions. Tiles containing less than 75% tissue content were discarded to minimise the inclusion of background or artefactual regions. Image preprocessing and patch extraction were performed using a Python-based pipeline leveraging OpenSlide for WSI handling, scikit-image and scipy for tissue masking, and OpenCV for image I/O operations. Patches meeting quality criteria were stored as PNG images in slide-specific directories for inspection, and were simultaneously saved in HDF5 datasets containing the raw image arrays and their corresponding tile coordinates. Metadata including slide identifier, native WSI resolution, number of retained patches, and extraction parameters were logged for each slide in a centralised CSV file to facilitate downstream model training and auditability.

### Stain normalization method

Patch-level color normalization was performed using Reinhard normalization, implemented via the HistomicsTK library. A predefined color distribution (target mean and standard deviation) derived from TCGA-A2-A3XS-DX1 (**2**) was used as the normalization reference.

(*2*) *Amgad, M. et al.* (*2019*)*. Structured crowdsourcing enables convolutional segmentation of histology images. Nature Communications, 10*(*1*), 1–13.

For each patch:

The image was converted from BGR to RGB format using OpenCV.

Reinhard normalization was applied using target mean [8.74, −0.12, 0.04] and target sigma [0.61, 0.11, 0.03].

The normalized image was clipped to valid pixel ranges, converted back to BGR, and saved in a parallel folder structure to the original dataset.

The normalization pipeline was executed using a custom Python script leveraging HistomicsTK, OpenCV, NumPy, and Pandas libraries.

All processed patches were saved as PNG images in a mirrored directory structure for downstream analysis.

### Feature extractor architecture (ResNet50, input-output dimensions)

Patch-level feature extraction was performed using ResNet50, a deep convolutional neural network with residual connections designed to facilitate gradient flow in deep architectures. The network was pre-trained on ImageNet and fine-tuned on the OSCC dataset by unfreezing the last 30 layers, while keeping earlier layers frozen to preserve generic image representations.

Input Patch Size: 256×256 pixels

Input Normalization: Mean = [0.485, 0.456, 0.406], Std = [0.229, 0.224, 0.225]

Batch Size: 600 patches per batch

Feature Output Dimension: 512-dimensional embedding for each patch

Processing Framework: PyTorch v2.5.1 on Python v3.12.3

Output Storage: Embeddings saved as pickle files for each WSI in class-organized directories.

### MIL model architecture, training strategy, hyperparameters

The developed AI pipeline employed a two-stage attention-based multiple instance learning (MIL) approach. Both stages utilized 512-dimensional patch-level embeddings extracted from a ResNet50 backbone network as inputs.

#### Stage 1: Binary MIL Model

To classify WSIs as either OSCC or normal tissue, an attention-based MIL classifier was developed. This model consisted of a linear projection followed by an attention mechanism. The attention module calculated learnable weights for each patch embedding, highlighting diagnostically relevant patches. Patch embeddings were aggregated using an attention-weighted pooling layer, followed by a fully connected layer and sigmoid activation function for binary classification.

#### Stage 2: Multiclass MIL Model

For OSCC subtype classification into well-differentiated (WD), moderately differentiated (MD), and poorly differentiated (PD) categories, an advanced attention-based MIL model was implemented, employing a Top-K pooling strategy. In this stage, the attention mechanism first calculated importance scores for patches. The model retained only the top-K patches with the highest attention weights (where K = 20, determined empirically), ensuring predictions were made based strictly on the most diagnostically informative regions. The selected top-K patch embeddings were then aggregated and passed through a fully connected classification layer with softmax activation for multiclass prediction.

#### Training Strategy

Both binary and multiclass MIL models shared a consistent training protocol, described below:

Cross-validation: Stratified three-fold cross-validation ensured balanced representation of all diagnostic categories across training and validation splits.

Loss Functions:

Binary Stage - Binary cross-entropy loss with logits (nn.BCEWithLogitsLoss), using class-specific weighting to mitigate imbalance.

Multiclass Stage - Cross-entropy loss (nn.CrossEntropyLoss), with class-specific weights derived from training data distribution.

Class Weighting: Class weights were computed per fold using the *sklearn.utils.class_weight.compute_class_weight* method to address inherent dataset imbalance.

Batch and Gradient Settings:

Batch Size: 1 WSI per batch due to variable patch counts across WSIs.

Gradient Accumulation: Gradients accumulated over 4 batches before parameter updates, stabilizing the training process.

Optimization and Learning Rate Scheduler:

Optimizer: Adam with an initial learning rate of 1e-4 and weight decay (L2 regularization) of 1e-3.

Scheduler: Cosine Annealing scheduler gradually reduced the learning rate from 1e-4 to 1e-6 over 50 epochs.

Epochs and Early Stopping: Training was conducted for up to 50 epochs, with early stopping applied based on validation loss to prevent overfitting.

Model Initialization: Kaiming initialization uniform for linear layers and normal for convolutional layers ensured stable convergence.

Computational Environment: Implemented using PyTorch v2.5.1 and Python v3.12.3, executed on GPU-enabled devices (CUDA).

#### Evaluation and Model Saving

At each epoch, model performance was evaluated using standard classification metrics: Accuracy, precision, recall, F1-score, Cohen’s kappa, and Matthews correlation coefficient (MCC). Detailed confusion matrices and classification reports were generated and saved separately for both training and validation datasets for each epoch. All trained model checkpoints, performance metrics, and logs were systematically archived per fold. The complete implementation is publicly accessible via our GitHub repository (https://github.com/NishaChaudhary23/oralpatho/).

### Metrics definitions (F1, recall, precision, AUC)

Performance was evaluated using precision, recall, F1-score, and area under the receiver operating characteristic curve (AUC-ROC). Precision was defined as the proportion of true positive predictions among all positive predictions, and recall as the proportion of true positives among all actual positives. The F1-score, calculated as the harmonic mean of precision and recall, was used to balance the trade-off between sensitivity and specificity. AUC-ROC was computed using the one-vs-rest strategy for multiclass settings. In addition, area under the precision–recall curve (AUC-PR) was computed to assess model performance under class imbalance. All metrics were computed using scikit-learn v1.4.2, with macro-averaged scores reported unless otherwise stated.

### Supplementary tables

**Table S1:** Model training hyperparameters value.

| Hyperparameter | Value |
| --- | --- |
| Cross-validation | Stratified three-fold |
| Optimizer | Adam |
| Initial learning rate | 1e-4 |
| Learning rate scheduler | Cosine Annealing (minimum LR: 1e-6) |
| Total epochs | 50 (early stopping enabled) |
| Top-K pooling (multiclass only) | K-20 (empirically determined) |
| Parameter initialization | Kaiming (uniform/normal) |

**Table S2:** Test performance metrics for binary and multiclass classification tasks. Performance values are averaged over three cross-validation folds for each task. Binary model was trained on OSCC vs. Normal classification; multiclass model predicted OSCC subtypes (WD, MD, PD).

| Metric | Binary (Mean ± SD) | Multiclass (Mean ± SD) |
| --- | --- | --- |
| Accuracy | 0.9091 ± 0.0124 | 0.6610 ± 0.0028 |
| Precision | 0.9095 ± 0.0118 | 0.6777 ± 0.0475 |
| Recall | 0.9091 ± 0.0124 | 0.5754 ± 0.0135 |
| F1 Score | 0.9086 ± 0.0128 | 0.6051 ± 0.0204 |
| Matthews Corr. Coeff. (MCC) | 0.8091 ± 0.0260 | 0.3862 ± 0.0076 |
| Cohen’s Kappa | 0.8077 ± 0.0274 | 0.3807 ± 0.0035 |

**Table S3:** Fold-wise per-class metrics for binary classification model.

| **Fold** | **Class** | **Precision** | **Recall** | **F1 Score** |
| --- | --- | --- | --- | --- |
| 1 | Normal | 0.92 | 0.88461538 | 0.90196078 |
|  | OSCC | 0.92682927 | 0.95 | 0.9382716 |
|  | accuracy | 0.92424242 | 0.92424242 | 0.92424242 |
|  | macro avg | 0.92341463 | 0.91730769 | 0.92011619 |
|  | weighted avg | 0.92413895 | 0.92424242 | 0.92396734 |
| 2 | Normal | 0.91304348 | 0.80769231 | 0.85714286 |
|  | OSCC | 0.88372093 | 0.95 | 0.91566265 |
|  | accuracy | 0.89393939 | 0.89393939 | 0.89393939 |
|  | macro avg | 0.8983822 | 0.87884615 | 0.88640275 |
|  | weighted avg | 0.89527224 | 0.89393939 | 0.8926094 |
| 3 | Normal | 0.88461538 | 0.88461538 | 0.88461538 |
|  | OSCC | 0.925 | 0.925 | 0.925 |
|  | accuracy | 0.90909091 | 0.90909091 | 0.90909091 |
|  | macro avg | 0.90480769 | 0.90480769 | 0.90480769 |
|  | weighted avg | 0.90909091 | 0.90909091 | 0.90909091 |

**Table S4:** Fold-wise per-class metrics for multiclass classification model.

| **Fold** | **Class** | **Precision** | **Recall** | **F1 Score** |
| --- | --- | --- | --- | --- |
| 1 | MD | 0.65079365 | 0.62121212 | 0.63565891 |
|  | PD | 0.63636364 | 0.33333333 | 0.4375 |
|  | WD | 0.66455696 | 0.73943662 | 0.7 |
|  | accuracy | 0.65762712 | 0.65762712 | 0.65762712 |
|  | macro avg | 0.65057142 | 0.56466069 | 0.59105297 |
|  | weighted avg | 0.65639148 | 0.65762712 | 0.65252365 |
| 2 | MD | 0.7029703 | 0.53787879 | 0.60944206 |
|  | PD | 0.9 | 0.42857143 | 0.58064516 |
|  | WD | 0.63043478 | 0.81690141 | 0.71165644 |
|  | accuracy | 0.66440678 | 0.66440678 | 0.66440678 |
|  | macro avg | 0.74446836 | 0.59445054 | 0.63391455 |
|  | weighted avg | 0.68208074 | 0.66440678 | 0.65659361 |
| 3 | MD | 0.67479675 | 0.62878788 | 0.65098039 |
|  | PD | 0.58333333 | 0.33333333 | 0.42424242 |
|  | WD | 0.65625 | 0.73943662 | 0.69536424 |
|  | accuracy | 0.66101695 | 0.66101695 | 0.66101695 |
|  | macro avg | 0.63812669 | 0.56718594 | 0.59019568 |
|  | weighted avg | 0.65935821 | 0.66101695 | 0.65620415 |

**Table S5:** Pathologist-guided evaluation of model interpretability on OSMF–OSCC WSIs.

| **Case ID** | **Mean Attention Score** | **% Region Match** | **Pathologist Comment** |
| --- | --- | --- | --- |
| Case 1 | 0.002 | 20% | Sparse tumor islands; some overlap with attention hotspots. |
| Case 2 | 0.0007 | 25% | Focal epithelial dysplasia detected; attention moderately aligned. |
| Case 3 | 0.001 | 25% | Tumor front partly captured; model missed peripheral spread. |
| Case 4 | 0.0004 | 45% | Good overlap with stromal invasion zones. |
| Case 5 | 0.0005 | 25% | Patchy detection near basal layers. |
| Case 6 | 0 | 75% | Correctly ignored fibrotic zones; true negatives. |
| Case 7 | 0 | 65% | Low attention but relevant benign epithelium well skipped. |
| Case 8 | 0 | 75% | Model silent on stromal areas; correct negative attention. |
| Case 9 | 0 | 25% | Limited detection; mostly false negative regions. |
| Case 10 | 0 | 50% | Weak signal; moderate region alignment with dysplastic epithelium. |

**SF1: Model performance metrics across three-fold cross-validation for binary and multiclass classifiers**

A. Performance metrics of the binary classifier trained using three-fold cross-validation. Each row shows training vs. validation accuracy (left), training vs. validation loss (middle), and bar plots of precision, recall, and F1-score for both train and validation sets (right).

B. Performance metrics of the multiclass classifier (WD, MD, PD) trained using three-fold cross-validation. Each row shows training vs. validation accuracy (left), training vs. validation loss (middle), and bar plots of precision, recall, and F1-score for both train and validation sets (right). The multiclass task exhibits increased variability and lower generalization compared to the binary classification task, highlighting the complexity of histological subtype separation.

**SF2 Model attention heatmaps over external test slides from institutional and public datasets.**

A. Representative examples from institutional OSCC samples show the original whole slide image (WSI) and the corresponding model-predicted regions with overlaid attention heatmaps.

B. Representative examples from the TCIA OSCC public dataset. High-attention regions predicted by the model are visualized, indicating spatial areas that contribute most to the diagnostic prediction. The heatmap color scale represents attention scores, with warmer colors (red) indicating high attention regions and cooler colors (blue) indicating lower contributions.

**SF3: Dataset demographics.**

A. Proportional distribution of Normal, OSCC, OSMF, and OSMF–OSCC samples from institutional and TCIA datasets.

B. Gender distribution in in-house samples, showing male predominance across all groups.

C. Age group breakdown by diagnosis; OSMF and OSMF-OSCC cohorts showed a higher prevalence in the 26-50 year age range.

D. Tobacco consumption habits reveal distinct patterns, with OSMF–OSCC patients reporting higher dual-use (chewing and smoking) compared to OSCC or OSMF alone.

E. Anatomical site distribution indicates buccal mucosa as the most common site in OSCC and OSMF-OSCC samples, with rare involvement of lip and palate.

**SF4: Model interpretability on OSMF-OSCC cases.**

Attention heatmaps for ten OSMF-OSCC slides, organized by descending mean attention score. Each column shows the original WSI and the corresponding model-predicted attention overlay. Case numbers indicate the percentage of tumor-relevant region overlap as estimated by a pathologist, followed by the mean attention score. While high-attention slides (e.g., Case 1, 3) show dense activation over basal epithelium and stroma, some low-attention slides (e.g., Case 6, 8) still align with histologically relevant regions, underscoring the model’s ability to detect subtle cues even under conservative activation.

**SF5: Boxplot showing the distribution of mean attention scores across all histological classes.**

Pairwise statistical significance was assessed using the two-sided Mann-Whitney U test. ‘n’ shows sample count. Comparisons are annotated within the plot using standard notations: **** (p < 0.0001), *** (p < 0.001), ** (p < 0.01), * (p < 0.05), and ns (not significant). Horizontal lines denote the pairwise comparisons. Attention scores tend to decrease across the continuum from normal to poorly differentiated OSCC. WDOSCC slides showed elevated attention variability, consistent with differentiated tumor architecture.

**SF6: Distribution and ranking of slide-level attention scores for OSMF–OSCC cases.**

A. Histogram of mean attention scores across all unseen OSMF-OSCC whole-slide images (WSIs) evaluated by the model. The distribution is highly right-skewed, with the majority of cases exhibiting near-zero attention, reflecting low model activation in response to subtle or ambiguous histological patterns.

B. Ranked barplot of OSMF-OSCC WSIs based on their mean attention scores. High-scoring cases (left) typically contain focal epithelial atypia or inflammation adjacent to tumor-prone zones, while low-scoring cases (right) are predominantly fibrotic or reactive with minimal dysplastic features. This disparity highlights the model’s capacity to discriminate morphologically relevant fields in pre-malignant conditions, despite the absence of oral carcinoma.

**SF7: Visual validation of WSI section-level expansion across representative slides.**

Multiple physically distinct tissue sections present on the same glass slide were treated as independent WSIs for model training and evaluation. This panel illustrates patch-level spatial clustering across selected slides using DBSCAN (Density-Based Spatial Clustering of Applications with Noise), confirming the presence of non-contiguous tissue sections that warranted independent processing (3). Each color denotes a unique section derived from the clustering of spatial coordinates.

(3) Ester M, Kriegel H-P, Sander J, Xu X. A density-based algorithm for discovering clusters in large spatial databases with noise. Proceedings of the 2nd International Conference on Knowledge Discovery and Data Mining (KDD); 1996. pp. 226–231.

**SF8: Web-based AI interactive interface for grading of oral cancer WSIs.**

A modular and interactive pipeline, *OralPatho*, developed for the AI-assisted grading of oral cancer histopathology WSIs. Left: Stepwise schematic of the diagnostic pipeline: starting from a raw histological WSI, the interface supports patch extraction, MIL-based binary OSCC detection (Stage 1), multiclass grading into WD, MD, PD (Stage 2), followed by slide-level prediction aggregation. Right: Interface screenshots illustrating user-accessible tabs for (i) patch generation and feature extraction, (ii) model inference for detection and grading, and (iii) visual inspection of attention heatmaps. The interface supports model customization and threshold adjustment to enable flexible and interpretable analysis.

