## Supplementary Figure 1 for "Dual-stage AI system for Pathologist-Free Tumor Detection and subtyping in Oral Squamous Cell Carcinoma"

### SF 1: Model performance metrics by binary and multiclass classifiers

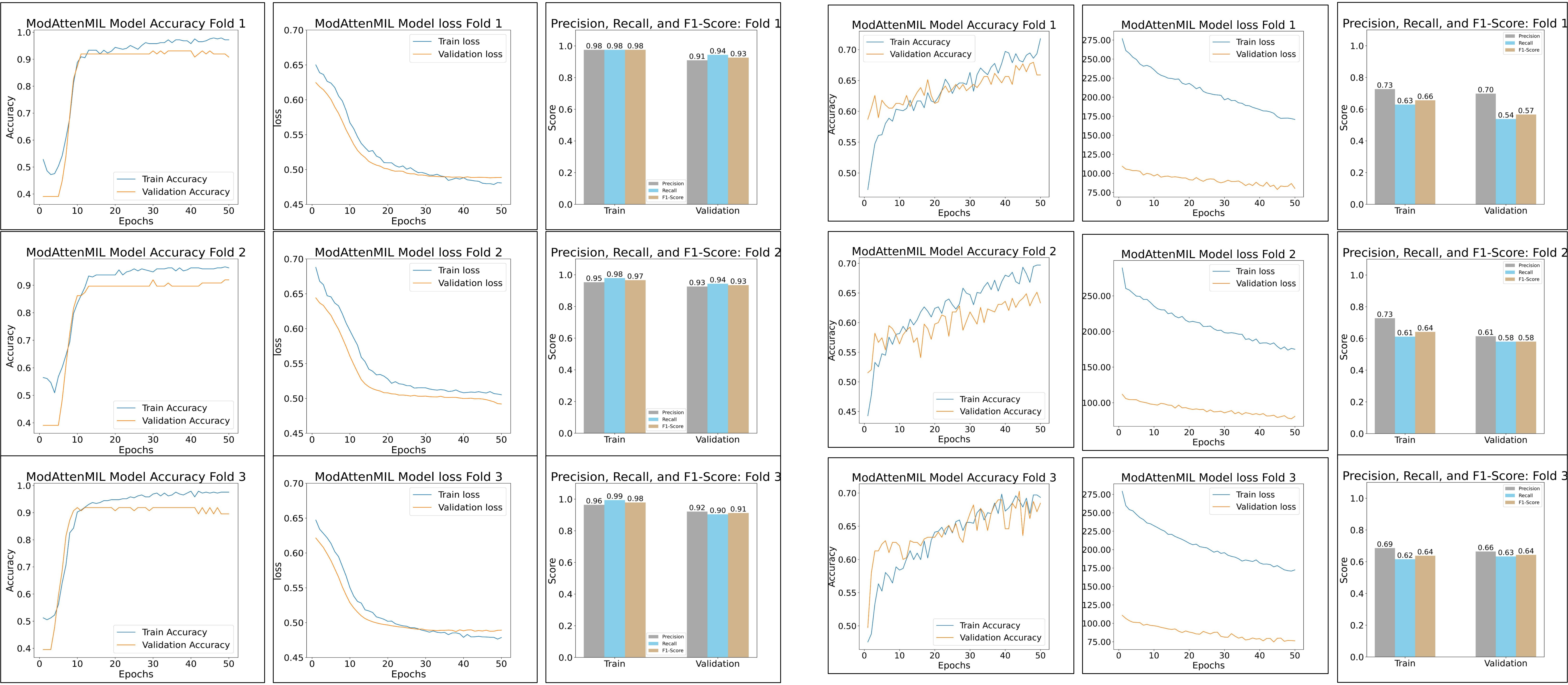


A Binary model performance

B Multiclass model performance
