## Supplementary figures and images for "Dual-stage AI system for Pathologist-Free Tumor Detection and subtyping in Oral Squamous Cell Carcinoma"

### Supplementary Figure 2

#### SF 2: Heatmap showing model prediction on external test data

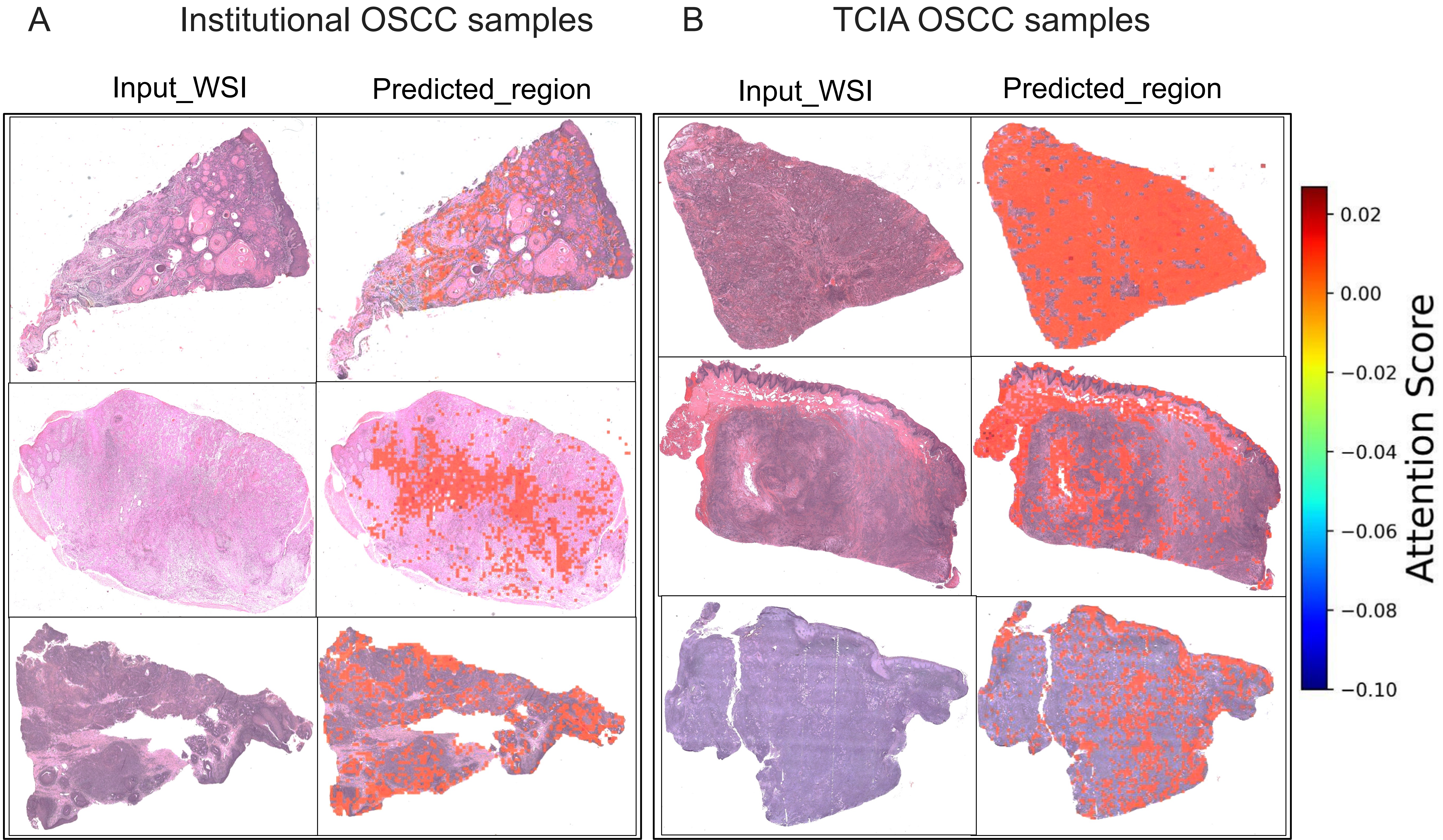

### Supplementary Figure 3

SF3: Dataset demographics.

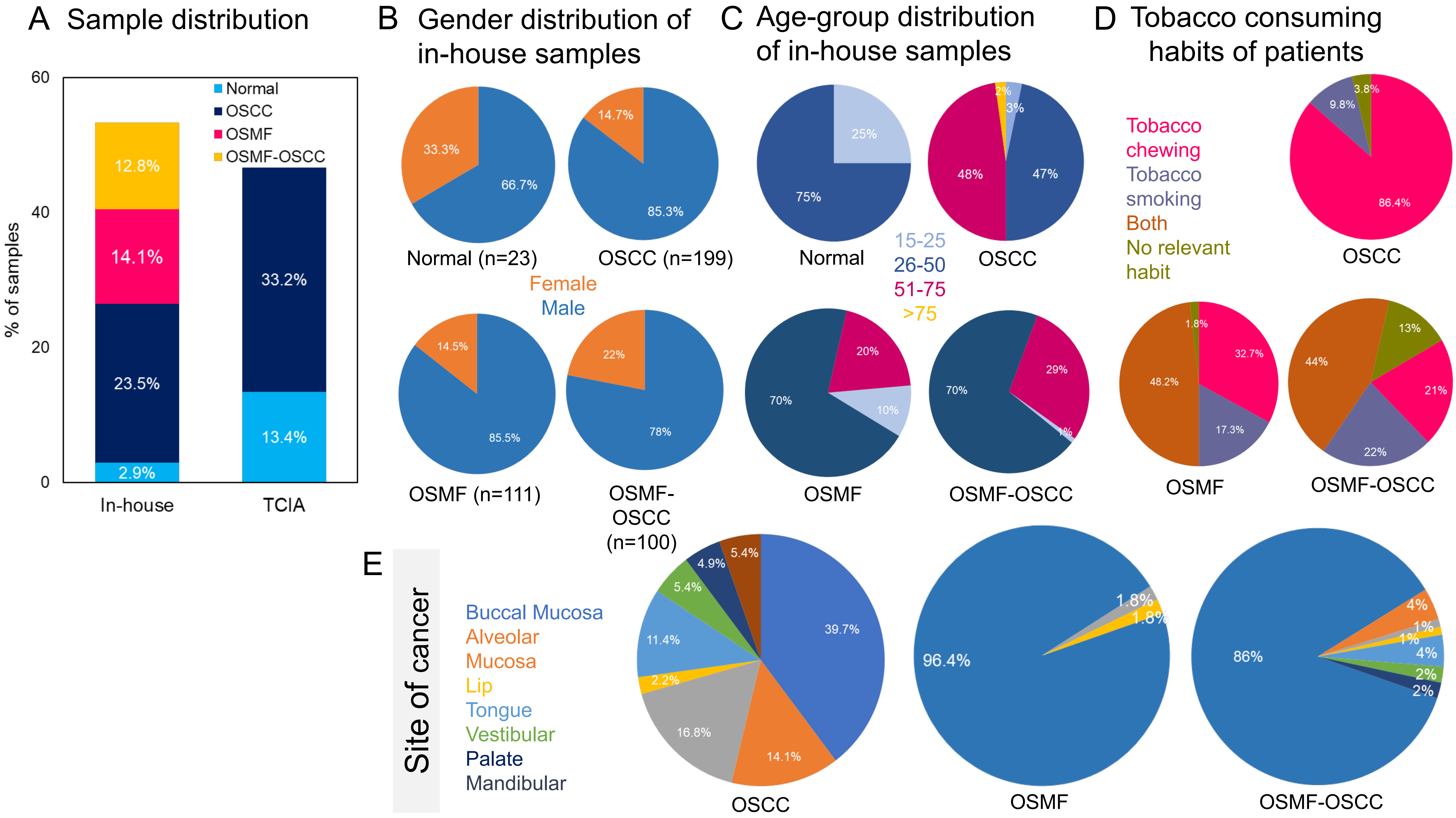

### Supplementary Figure 4

#### SF4: Model interpretability on OSMF-OSCC cases.

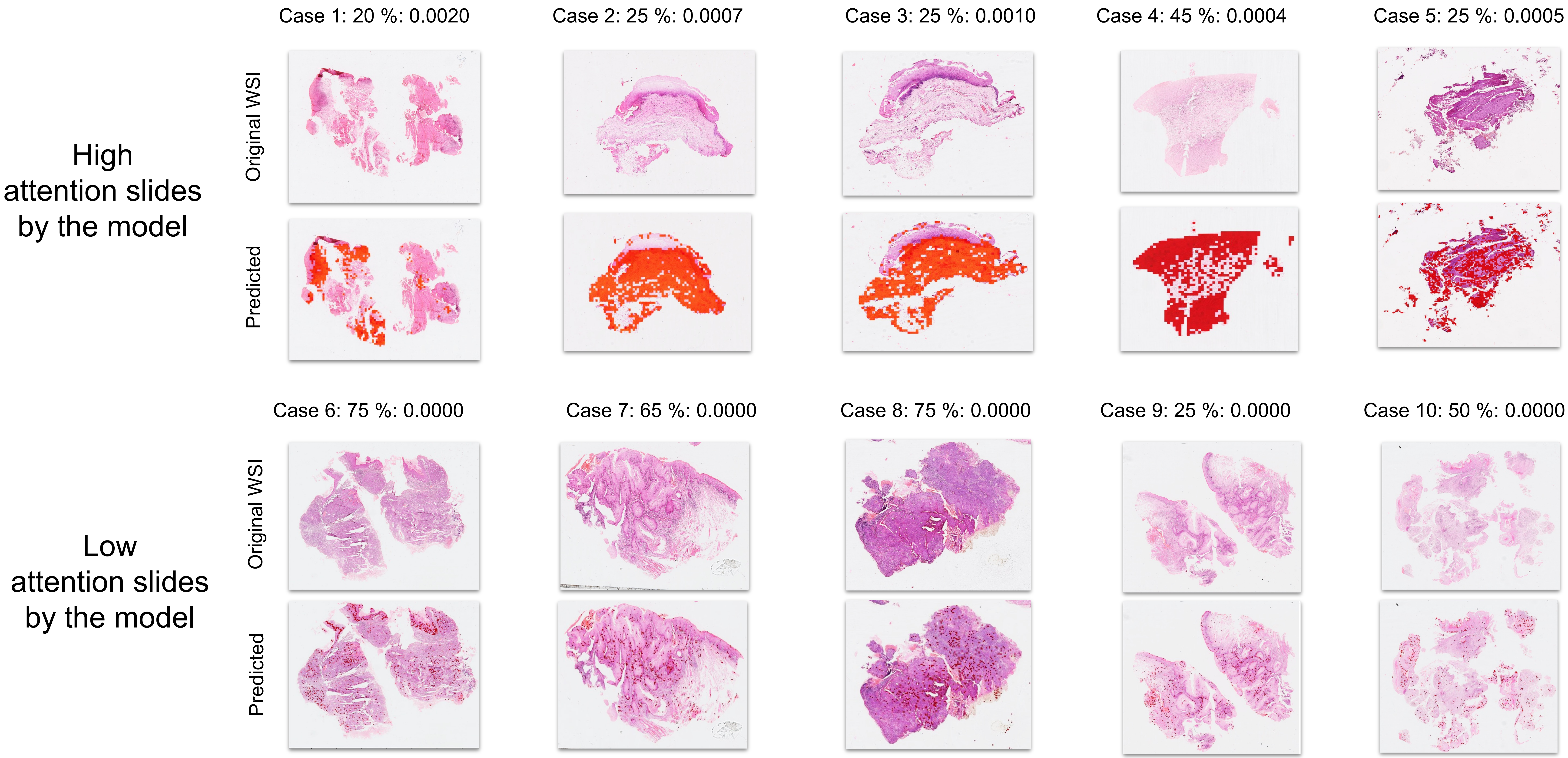

### Supplementary Figure 6

SF6: Distribution and ranking of slide-level attention scores for OSMF–OSCC cases.

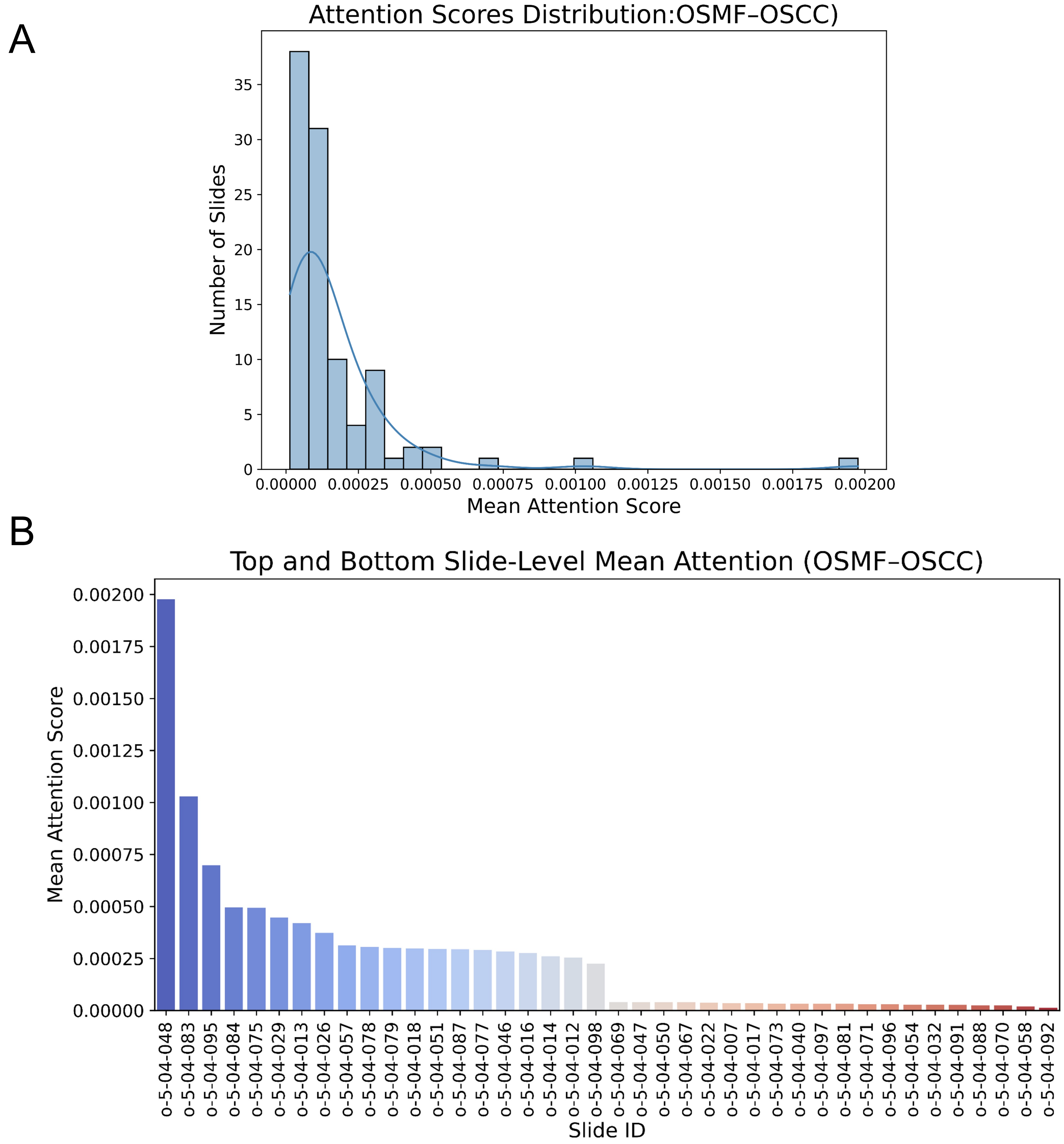

### Supplementary Figure 7

#### SF7: Visual validation of WSI section-level expansion across representative slides.

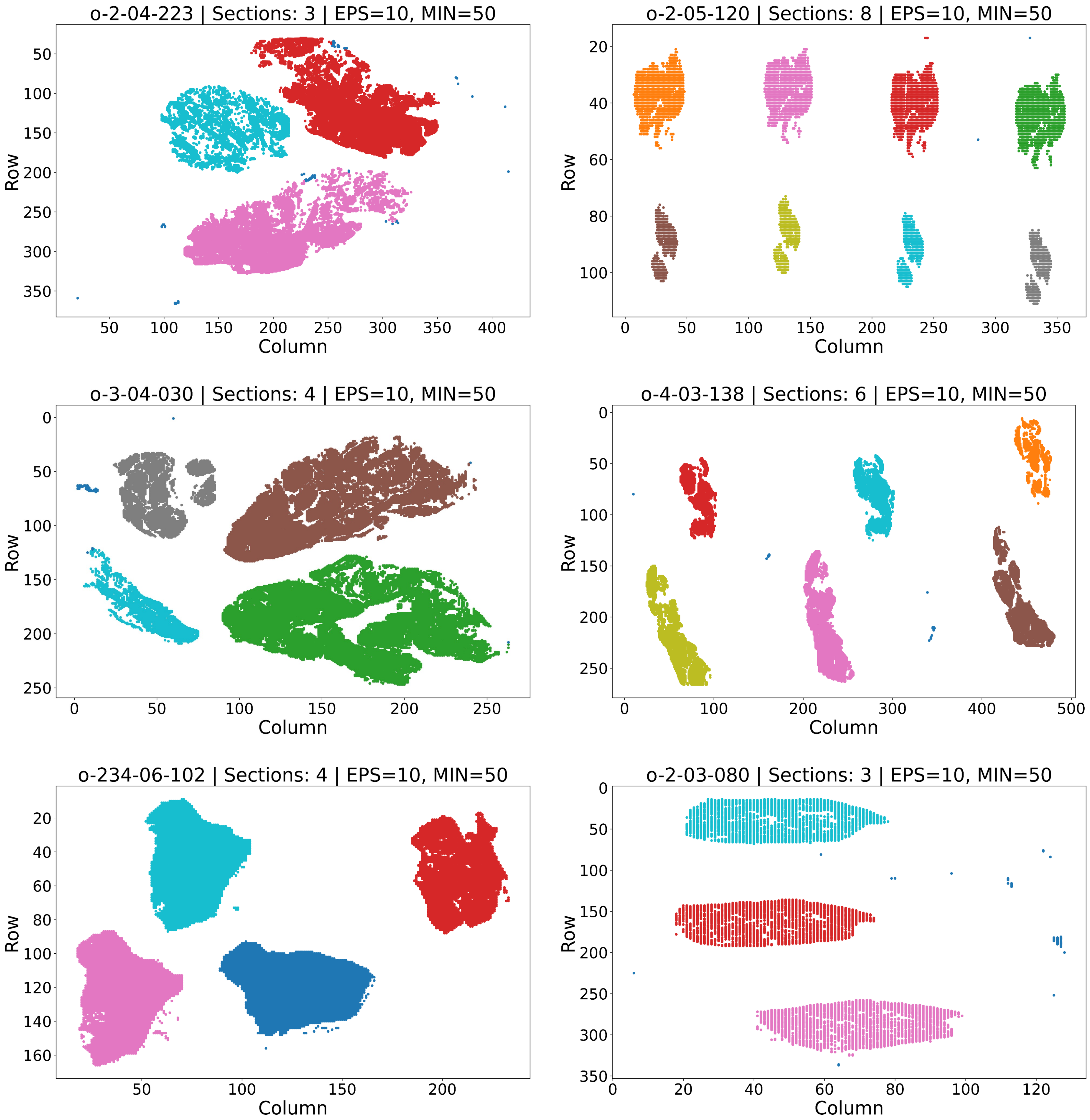

### Supplementary Figure 8

#### SF8: Web-based AI interactive interface for grading of the oral WSI.

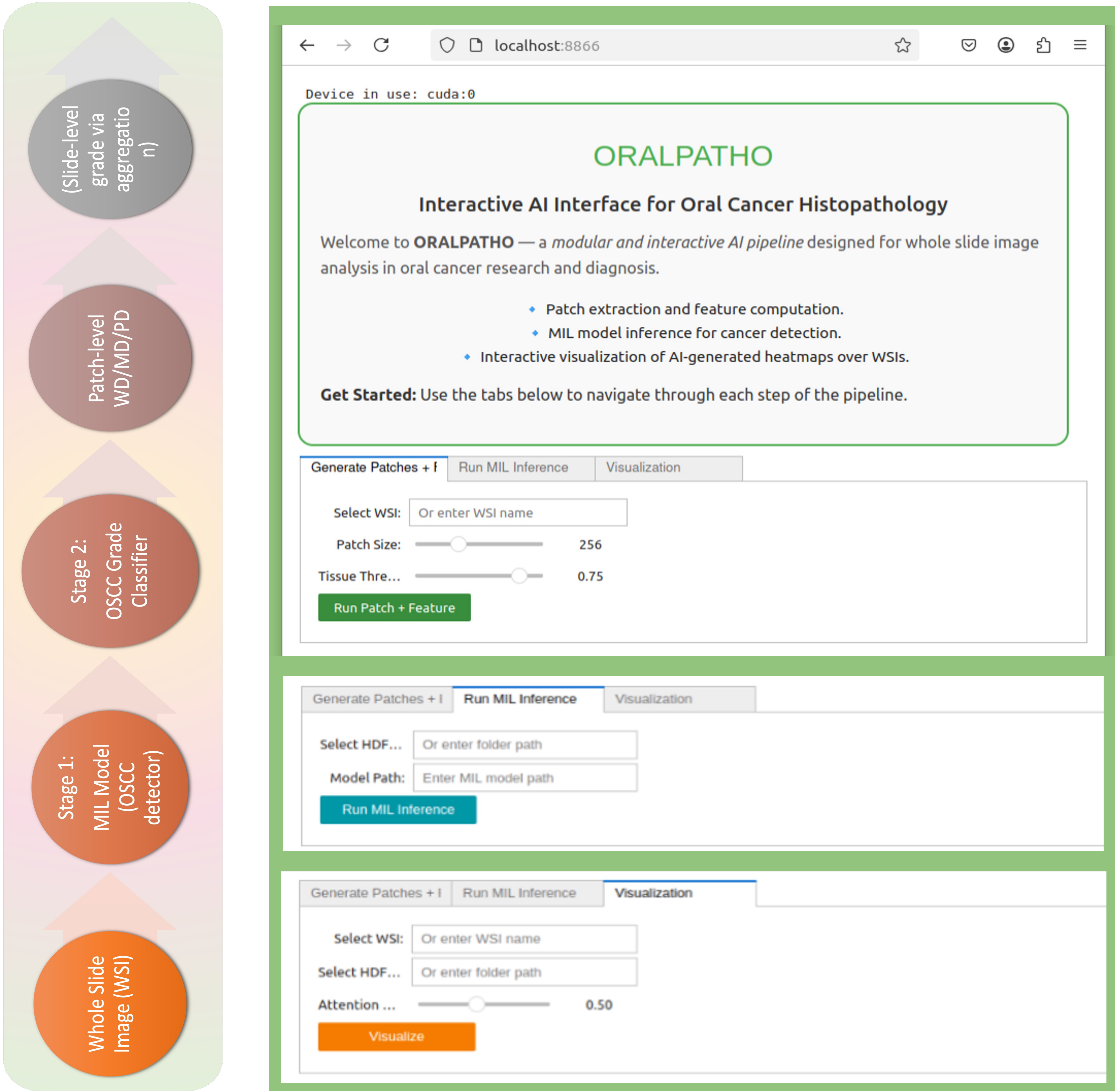
