## Supplementary Figure 5 for "Dual-stage AI system for Pathologist-Free Tumor Detection and subtyping in Oral Squamous Cell Carcinoma"

SF5: Boxplot showing the distribution of mean attention scores across all histological classes.

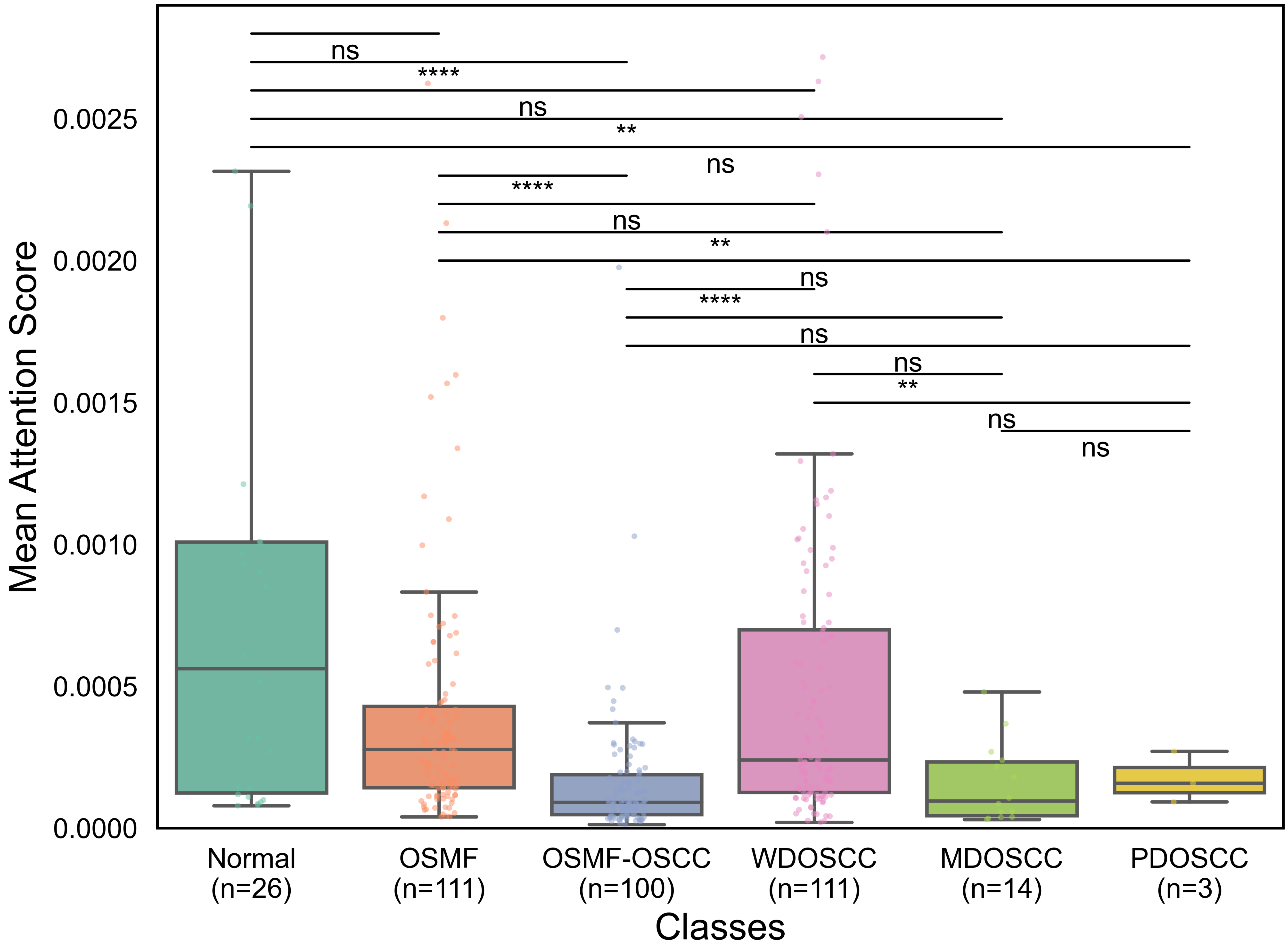
